# COVID-19 in healthcare workers in three hospitals in the South of the Netherlands, March 2020

**DOI:** 10.1101/2020.04.26.20079418

**Authors:** Reina S. Sikkema, Suzan Pas, David F. Nieuwenhuijse, Áine O’Toole, Jaco Verweij, Anne van der Linden, Irina Chestakova, Claudia Schapendonk, Mark Pronk, Pascal Lexmond, Theo Bestebroer, Ronald J. Overmars, Stefan van Nieuwkoop, Wouter van den Bijllaardt, Robbert G. Bentvelsen, Miranda M.L. van Rijen, Anton G.M. Buiting, Anne J.G. van Oudheusden, Bram M. Diederen, Anneke M.C. Bergmans, Annemiek van der Eijk, Richard Molenkamp, Andrew Rambaut, Aura Timen, Jan A.J.W. Kluytmans, Bas B. Oude Munnink, Marjolein F.Q. Kluytmans van den Bergh, Marion P.G. Koopmans

## Abstract

Ten days after the first reported case of SARS-CoV-2 infection in the Netherlands, 3.9% of healthcare workers (HCWs) in nine hospitals located in the South of the Netherlands tested positive for SARS-CoV-2 RNA. The extent of nosocomial transmission that contributed to the HCW infections was unknown.

We combined epidemiological data, collected by means of structured interviews of HCWs, with whole genome sequencing (WGS) of SARS-CoV-2 in clinical samples from HCWs and patients in three of nine hospitals that participated in the HCW screening, to perform an in-depth analysis of sources and modes of transmission of SARS -CoV-2 in HCWs and patients.

A total of 1,796 out of 12,022 HCWs (15%) of the three participating hospitals were screened, based on clinical symptoms, of whom 96 (5%) tested positive for SARS-CoV-2. We obtained complete genome sequences of 50 HCWs and 18 patients. Most sequences grouped in 3 clusters, with 2 clusters displaying local circulation within the region. The observed patterns are most consistent with multiple introductions into the hospitals through community acquired infections, and local amplification in the community.

Although direct transmission in the hospitals cannot be ruled out, the data does not support widespread nosocomial transmission as source of infection in patients or healthcare workers.

## Introduction

Early December 2019 an outbreak caused by a novel coronavirus, linked to a seafood and live animal wholesale market in Wuhan, China was discovered^1^. The clinical spectrum of disease varies from asymptomatic or mild symptomatic infections to severe respiratory symptoms and mortality, with older age groups generally presenting with more severe disease and higher death rates ^2,3^. Since its discovery, SARS-CoV-2 has rapidly spread across the globe. On April 22nd a total of 177 countries reported SARS-CoV-2, adding up to over 2,590,000 reported cases and more than 179,000 deaths worldwide^4^.

Healthcare workers (HCWs) are at greater risk of being exposed to SARS-CoV-2 and could potentially play a role in hospital outbreaks. Nosocomial outbreaks of SARS-CoV and MERS-CoV are thought to have played a crucial role in the amplification and spread of these viruses. For MERS-CoV, hospital outbreaks have been responsible for approximately 50% of confirmed human cases, of which around 40% were healthcare workers^5,6^. Currently the extent of SARS-CoV-2 transmission in health care settings is unclear, as well as risk factors associated with infection in health care settings. During the WHO-China Joint Mission on coronavirus disease 2019 (COVID-19) 2,055 laboratory-confirmed healthcare workers from 476 hospitals were reported, mostly from Hubei province (88%). Most HCW infections were thought to have been infected within household settings rather than in a health care setting, although conclusive evidence was lacking^3^.

On February 27, the first Dutch patient tested positive for SARS-CoV-2 RNA after a holiday to Lombardy, Italy ^7^. In the following week, the number of cases increased to 128, with an increasing proportion of cases without a known source of infection. These cases included nine HCWs from two hospitals in North Brabant, in the South of the Netherlands ^2,8^. Therefore, the Dutch national Outbreak Management Team advised to extend this screening of HCWs to other hospitals in the province of North Brabant, to assess possible community transmission. From March 6^th^ to March 8^th^, 1,097 employees of nine hospitals were tested of which 3.9% was found to be positive for SARS-CoV-2 ^8^. A follow up study was done at three hospitals to assess the clinical presentations of COVID-19 of these HCWs^2^, and in the present study an in-depth investigation was done to understand sources and modes of transmission. The scarcity of personal protective equipment led to changes in policy during the initial phases of the outbreak response, also triggering a debate on possible risks to HCW.

Here, we combined epidemiological data with whole genome sequencing of SARS-CoV-2 in clinical samples from HCWs and patients in three different hospitals to perform an in-depth analysis of sources and modes of transmission of SARS -CoV-2 in HCWs and patients in the same hospitals.

## Material and methods

### Sample collection

A cross-sectional study was conducted in two teaching hospitals (700-bed Amphia Hospital, Breda, the Netherlands; 800-bed Elisabeth-TweeSteden Hospital, Tilburg, the Netherlands, the Netherlands) and one regional hospital (600-bed Bravis Hospital, Roosendaal and Bergen op Zoom), employing a total of 12,022 HCWs. Between March 2, 2020 and March 12, 2020, HCWs in these three hospitals who experienced fever or mild respiratory symptoms in the last ten days were voluntarily tested for SARS-CoV-2 infection using oro-/nasalpharyngeal swabs in universal transport medium or E-swab medium (Copan, Italy), following the local infection control policy during outbreaks.

Structured interviews were conducted to document history of foreign travel and attendance of public events with more than 50 persons, such as the yearly carnival in February, for all HCWs with confirmed SARS-CoV-2 infection (Annex 2 - questionnaire). Ethical approval was obtained from the Ethics Committee Brabant, with a waiver of written informed consent (METC Brabant/20.134/NW2020–26). Verbal informed consent was obtained from all HCWs for SARS-CoV-2 testing, sequencing and data collection. Data was de-identified before analysis.

### RT-PCR

In the Bravis and Amphia hospital, total nucleic acids were extracted after an external lysis step (1:1 with lysis/binding buffer (Roche Diagnostics, the Netherlands), using MagnaPure96 (Roche) with an input volume of 500 μl and output volume of 100 μl. The extraction was internally controlled by the addition of a known concentration of phocine distemper virus (PDV)^9^. Subsequently 10 μl extracted nucleic acids was amplified in three singleplex reactions in 25 μl final volume, using TaqMan Fast Virus 1-Step Master Mix (Thermofisher, Nieuwerkerk a/d IJssel, the Netherlands), and 1 μl of primers and probe mixture for E gene, RdRp gene and PDV^10^. Amplification was performed in a 7500SDS (Thermofisher) machine with a cycling profile of 5 min at 50°C, 20 s at 95°C, 45 cycles of 3 s at 95°C and 30 s at 58°C. Alternatively, in ETZ hospital, total nucleic acids were extracted, with a known concentration of PDV as internal control, using the QIAsymphony DSP virus/pathogen midi kit and pathogen complex 400 protocol of the QIAsymphony Sample Processing (SP) system (Qiagen, Hilden, Germany) with an input volume 400 μl and output volume of 110 μl. Amplification reactions were performed in a volume of 25 μL with TaqMan® Fast Virus 1-Step Master Mix (Thermofisher) and 10 μL extracted nucleic acids. A duplex PCR for E-gen/PDV ^10,11^ with optimized primer and probe concentrations were performed. Amplification using Rotorgene (QIAgen) consisted of 5 min at 50°C, 15 min at 95°C followed by 45 cycles of 15 s at 95°C, 30 s at 60°C, and 15s at 72°C. Validations of RT-PCR procedures were performed according to International Standards Organization guidelines 15189 (http://www.iso.org/iso/search.htm).

### Whole genome sequencing

Samples were selected based on cycle threshold (Ct) value (<32). A SARS-CoV-2 specific multiplex PCR for Nanopore sequencing was performed, similar to amplicon-based approaches as previously described^12,13^. The resulting raw sequence data was demultiplexed using qcat (https://github.com/nanoporetech/qcat). Primers were trimmed using cutadapt^14^ after which a reference based alignment was performed using minimap2^15^ to GISAID sequence EPI_ISL_412973. The consensus genome was extracted and positions with a coverage <30 were replaced with an “N” with a custom script using biopython and pysam, as previously described^12^. Mutations in the genome were confirmed by manually checking the alignment and homopolymeric regions were manually checked and resolved consulting the reference genome. Complete whole genomes were defined as having >90% genome coverage.

### Sequence data analysis

All available full-length SARS-CoV-2 genomes were retrieved from GISAID^16^ on March 20 2020 and aligned with the newly obtained SARS-CoV-2 sequences in this study using MUSCLE^17^. Sequences with >10% “Ns” were excluded. The alignment was manually checked for discrepancies after which IQ-TREE^18^ was used to perform a maximum likelihood phylogenetic analysis with the GTR+F+I+G4 model as best predicted model. The ultrafast bootstrap option was used with 1,000 replicates.

Clusters 1 (B2), 2 (B) and 3 (B1) were determined based on visual clustering as well as lineage designations as proposed by Rambaut et al^19^.

### Minimum spanning tree visualization

The same alignment as for the maximum likelihood phylogenetic analysis was used to create a minimum spanning tree (MST). The code to generate the MST was written in the R programming language. The ape^20^ and igraph^21^ software packages were used to write the code to generate the minimum spanning tree and the visNetwork (https://github.com/datastorm-open/visNetwork) software package was used to generate the visualization. Pairwise sequence distance, used to generate the network, was calculated by adding up the absolute nucleotide distance and indel-block distance. Unambiguous positions were dealt with in a pairwise manner. Sequences that were mistakenly identified as identical, due to transient connections with sequences containing missing data were resolved.

### BEAST/ML trees

The multiple sequence alignment was curated and any error-rich sequences or sequences without a date were removed. The alignment was manually inspected and trimmed of the 5’ and 3’ UTRs in Geneious v11.1.3 (geneious.com) to include only coding regions. The final length of the alignment was 29,408 nucleotides. Bayesian phylogenetic trees were estimated using BEAST v1.10.4^22^ using an HKY nucleotide subsititution model^23^ and a strict molecular clock. Two independent chains were run for 100 million states, with a skygrid coalescent prior^24,25^. Trees and parameters were sampled every 10,000 states.

LogCombiner was used to combine the independent chains and to remove the burn-in from the tree file and Tracer was used to assess convergence^26^. The maximum clade credibility (MCC) tree was inferred using TreeAnnotator and visualized using baltic (https://github.com/evogytis/baltic) and custom python scripts.

### Statistical analysis

Epidemiological data, as collected by the structured interviews, were entered in CASTOR EDC (version 2019; https://castoredc.com). Continuous variables were expressed as medians and ranges. Categorical variables were summarized as counts and percentages. All analyses were performed with SPSS version 25.0 (IBM, Armonk, NY, USA). Differences between clusters were expressed in a similar manner. Due to the nature of the data and the low numbers of sequences per cluster, statistical analysis to compare clusters was not performed.

## Results

A total of 1,796 out of 12,022 HCWs (15%) of the three participating hospitals were screened, of whom 96 (5%) tested positive for SARS-CoV-2 RNA. At the Amphia Hospital, 42 (5%) of 783 HCWs tested positive, at the ETZ Hospital 44 (8%) of 570 HCWs and at the Bravis Hospital 10 (2%) of 443 HCWs. HCWs with COVID-19 were employed in 58 different departments, including 42 medical wards. Their median age was 49 years (range 22–66 years) and 17% (16/96) were male (Table 1). Twenty staff members without direct patient contact tested positive for SARS-CoV-2 RNA, six of whom (30%) reported contact with SARS-CoV-2 RNA positive colleagues. Ten HCWs reported a history of foreign travel in the 14 days prior to the onset of symptoms, three (3%) of whom had travelled to Northern-Italy. Sixty (63%) HCWs had celebrated carnival in the 14 days prior to the onset of symptoms, mostly in Tilburg, Breda and Prinsenbeek. One HCW, with February 21 as the day of first symptoms, reported to have attended several carnival events whilst being symptomatic, but unaware of having COVID-19. Thirty-one (32%) HCWs reported close contact with an individual known with COVID-19 in the 14 days prior to the onset of symptoms, being a patient (n=3), a colleague (n=18), a household member (n=1) or another person outside the hospital (n=9).

**Table 1.**
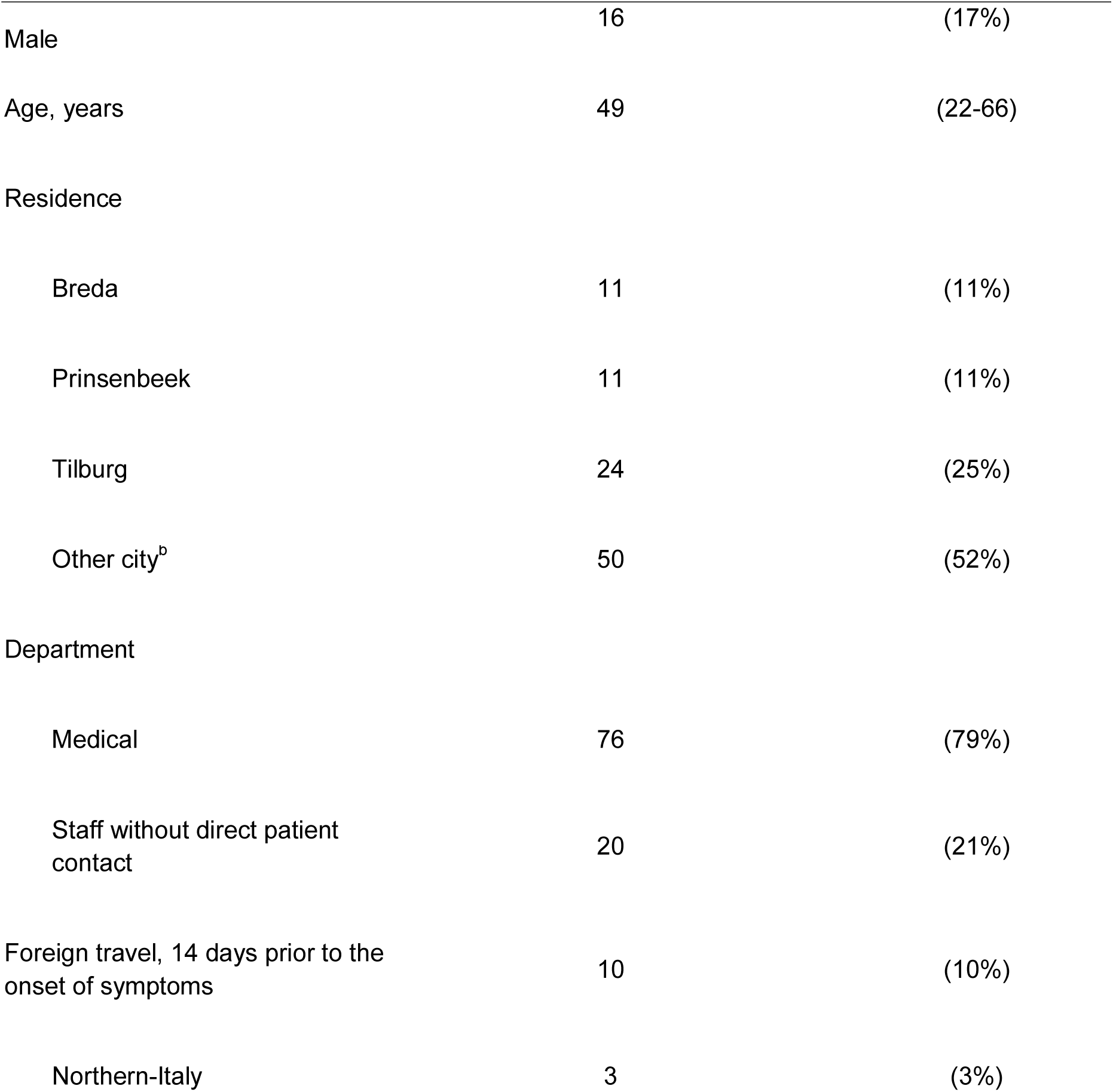
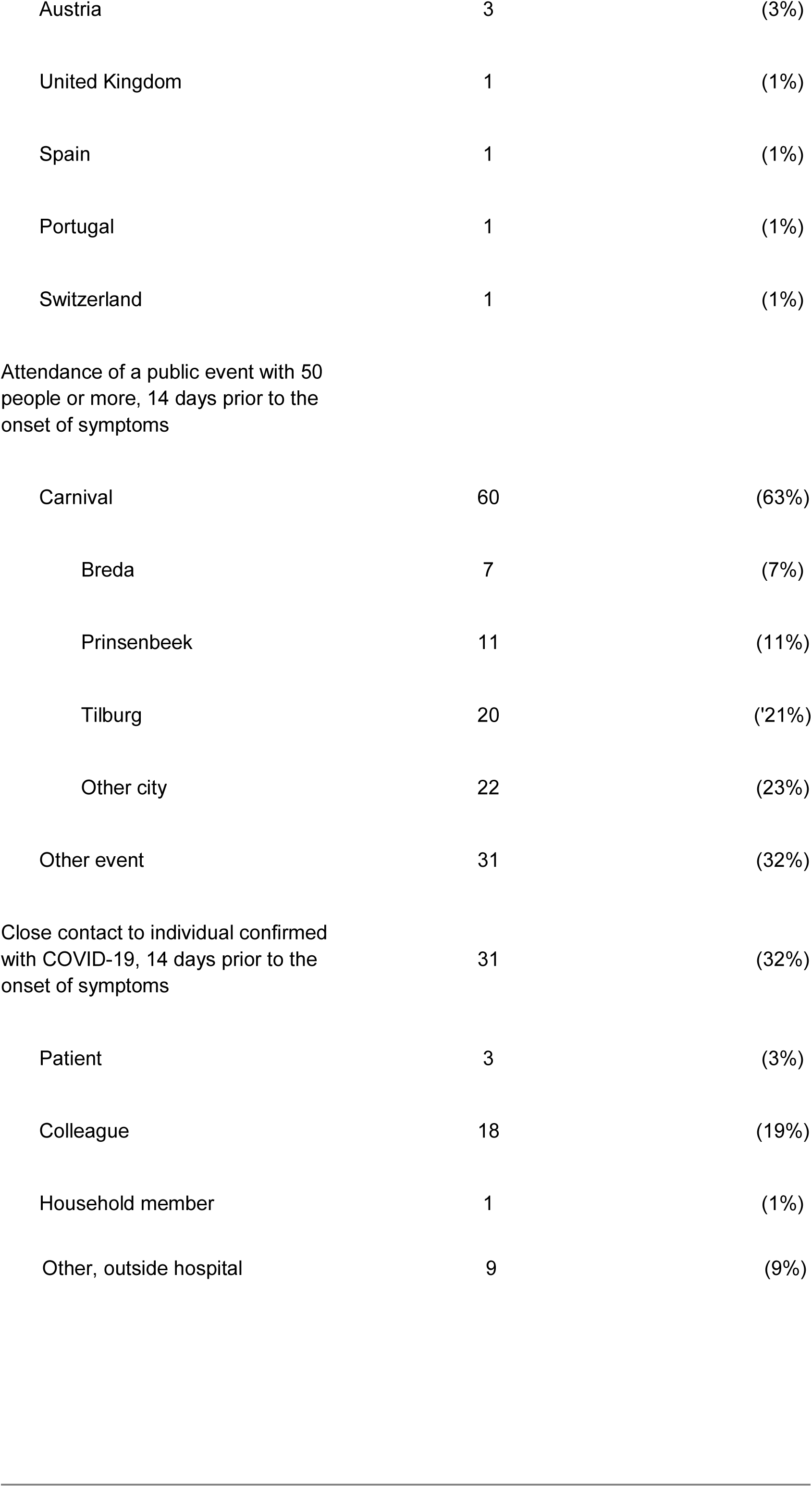
Descriptive characteristics of 96 healthcare-workers with COVID-2019 in the Netherlands in March 2020.

We obtained complete genome sequences of 50 HCWs (13 ETZ, 31 Amphia, 6 Bravis) of samples with sufficient viral loads. In addition we obtained sequences from five patients at the ETZ hospital (samples taken between February 27 - March 3, 2020), nine patients from the Amphia hospital (samples taken between March 5 and March 9 2020) and four patients from the Bravis hospital (March 8 and March, 2020).

Most HCW sequences in this study (46/50; 92%) grouped in three clusters (Figure 1A; Annex 1). Cluster 1 contains 21 out of 68 sequences in this study, ten of which are identical. 90% (19/21) of these sequences originated from HCWs and patients of the Amphia hospital, compared to 41% of the sequences in cluster 2 (14/34). HCWs in cluster 1 were associated with Prinsenbeek and Breda, either by carnival or residence, more often compared to the other clusters (Figure 1A; Figure 1B; Table 2). Cluster 2 contained 34 sequences including 19 identical sequences, of HCWs from all three hospitals, including 85% of ETZ hospital HCWs, unlike what was observed for cluster 1 (Figure 1A; Figure 1B; Table 2). The third cluster contained a small part of HCWs from this study, but reflects a larger cluster of sequences from persons with travel history to Italy, as described elsewhere^13^. However, only two of six HCWs in this cluster reported to have traveled to either Italy or Austria.

**Table 2.**
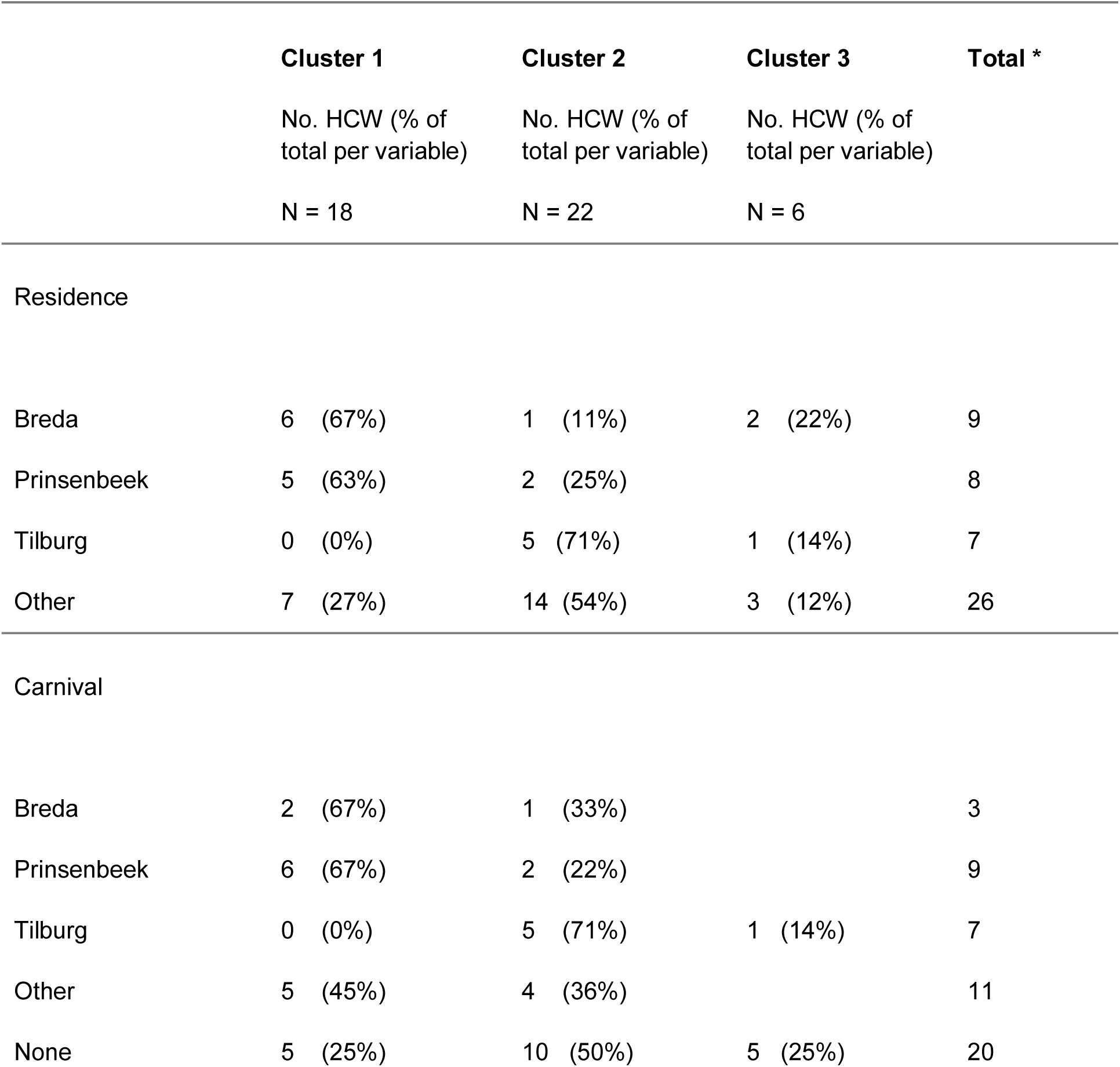
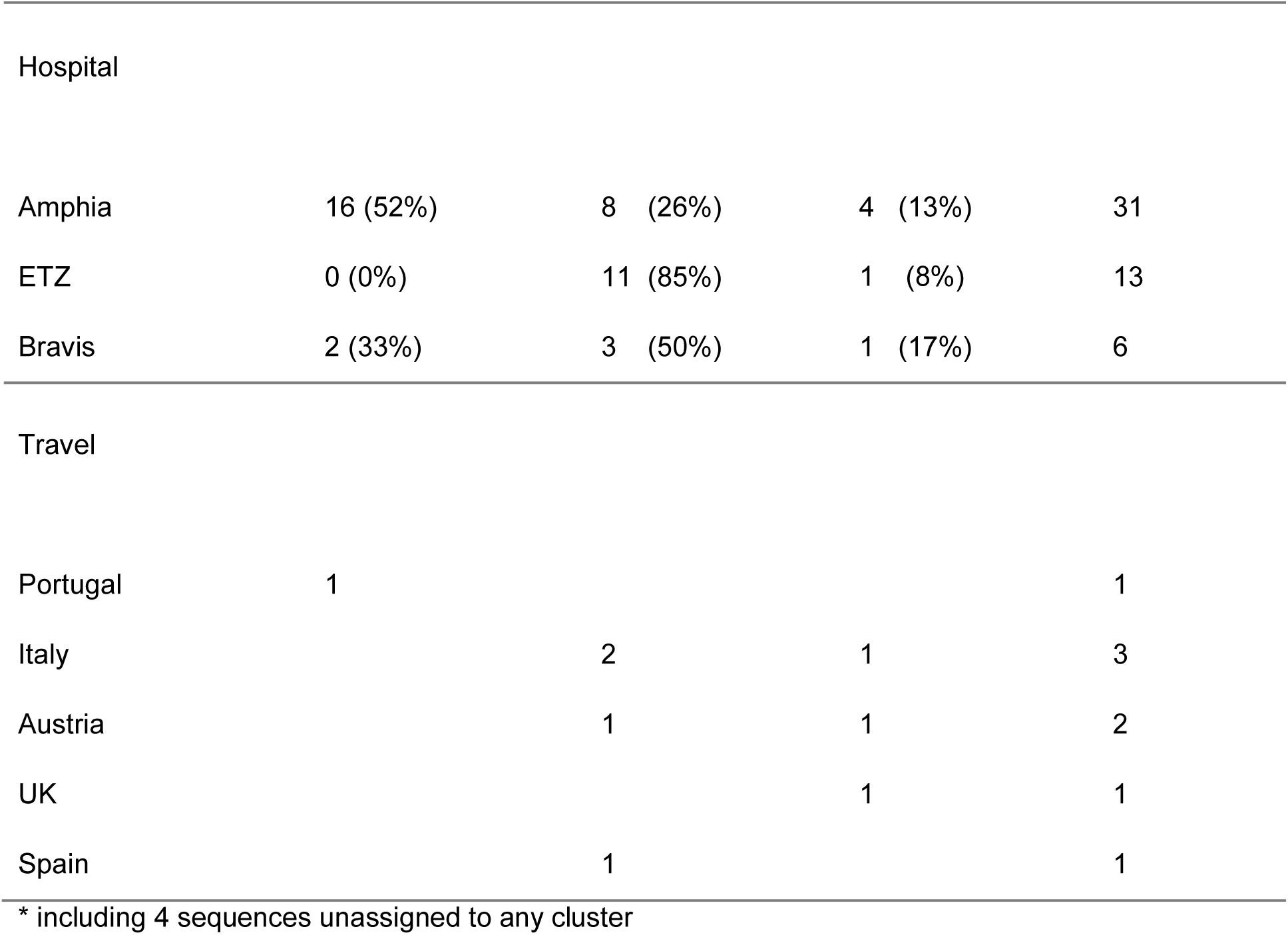
Descriptive characteristics of 3 clusters with 46 healthcare-workers with coronavirus disease 2019 in the Netherlands in March 2020.

|  | **Cluster 1** | **Cluster 2** | **Cluster 3** | **Total \*** |
| --- | --- | --- | --- | --- |
|  | No. HCW (% of total per variable) | No. HCW (% of total per variable) | No. HCW (% of total per variable) |  |
|  | N = 18 | N = 22 | N = 6 |  |
| Residence |  |  |  |  |
| Breda | 6 (67%) | 1 (11%) | 2 (22%) | 9 |
| Prinsenbeek | 5 (63%) | 2 (25%) |  | 8 |
| Tilburg | 0 (0%) | 5 (71%) | 1 (14%) | 7 |
| Other | 7 (27%) | 14 (54%) | 3 (12%) | 26 |
| Carnival |  |  |  |  |
| Breda | 2 (67%) | 1 (33%) |  | 3 |
| Prinsenbeek | 6 (67%) | 2 (22%) |  | 9 |
| Tilburg | 0 (0%) | 5 (71%) | 1 (14%) | 7 |
| Other | 5 (45%) | 4 (36%) |  | 11 |
| None | 5 (25%) | 10 (50%) | 5 (25%) | 20 |
| Hospital |  |  |  |  |
| Amphia | 16 (52%) | 8 (26%) | 4 (13%) | 31 |
| ETZ | 0 (0%) | 11 (85%) | 1 (8%) | 13 |
| Bravis | 2 (33%) | 3 (50%) | 1 (17%) | 6 |
| Travel |  |  |  |  |
| Portugal | 1 |  |  | 1 |
| Italy |  | 2 | 1 | 3 |
| Austria |  | 1 | 1 | 2 |
| UK |  |  | 1 | 1 |
| Spain |  | 1 |  | 1 |

\* including 4 sequences unassigned to any cluster

**Figure 1A:**
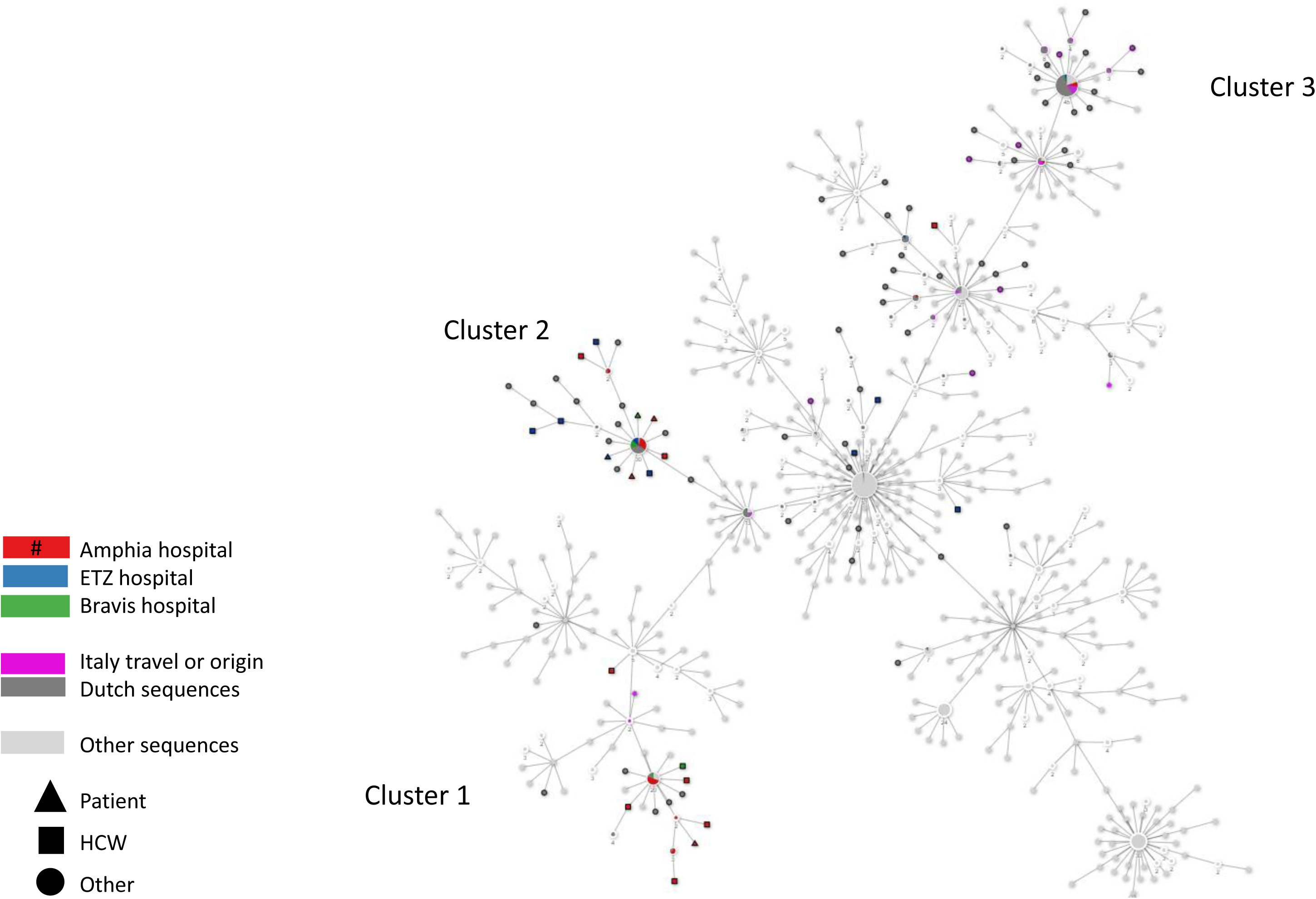
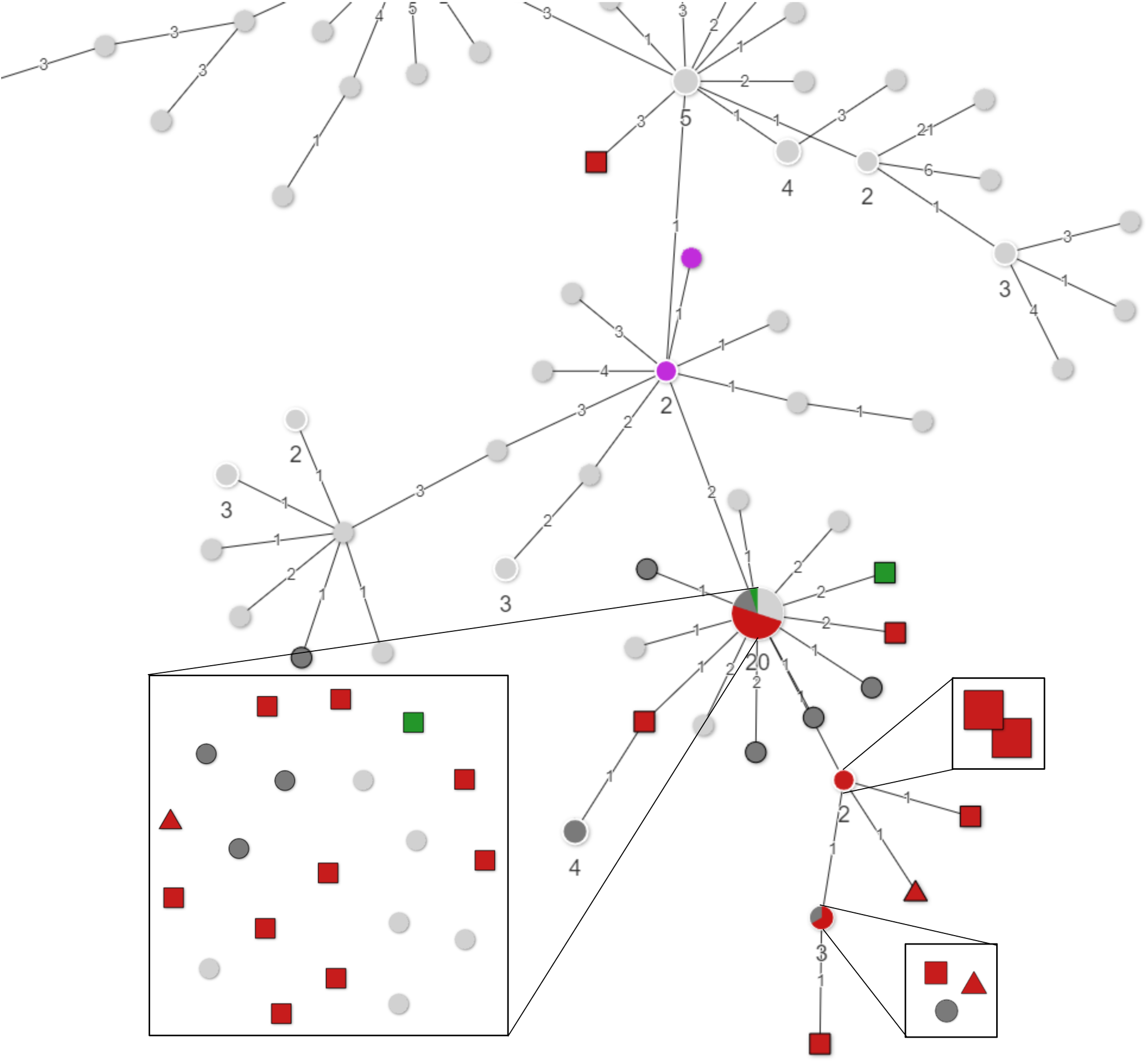
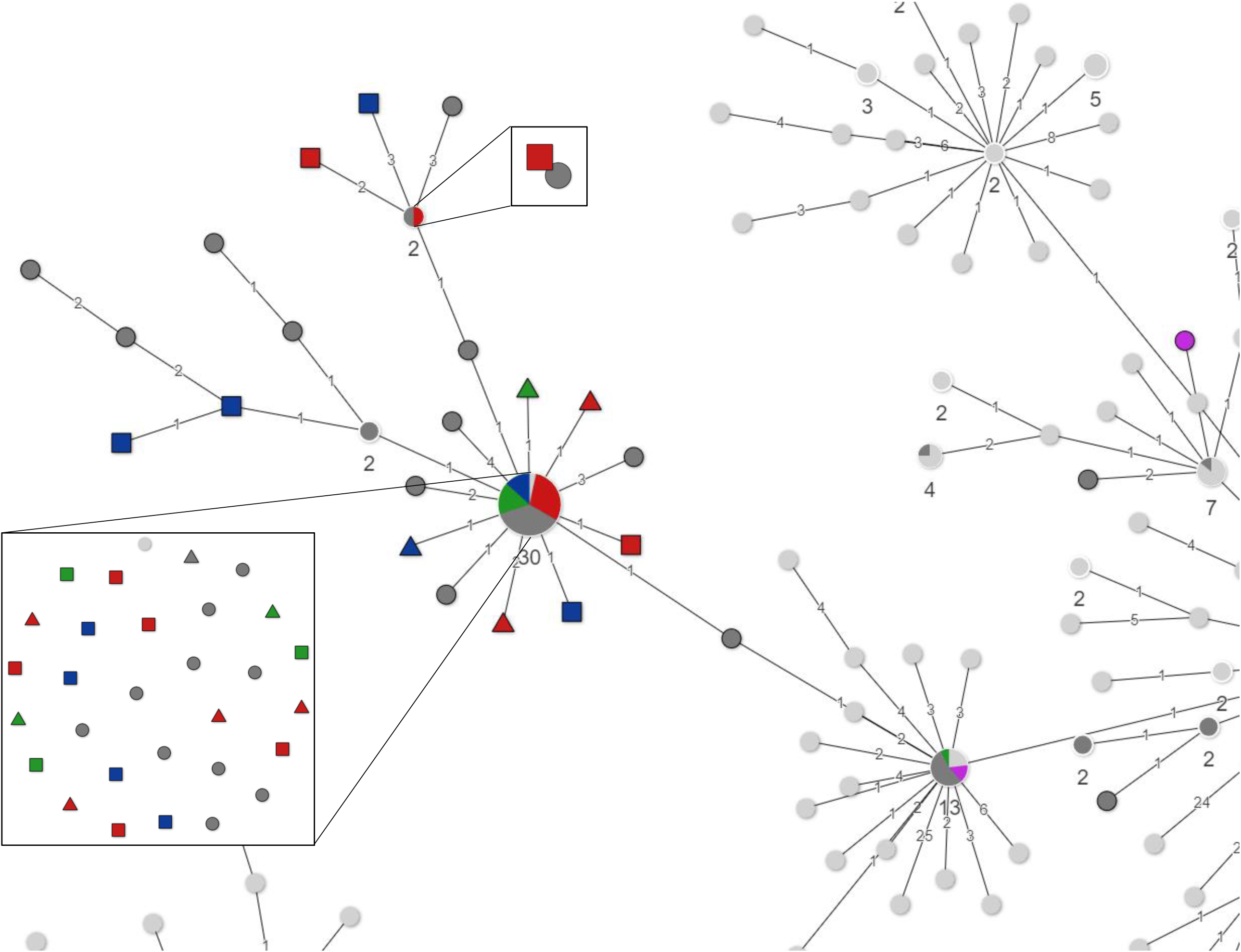
Minimum Spanning Tree of all available full length SARS-CoV-2 genomes retrieved from GISAID on March 20 2020, showing three clusters of HCW and patient sequences from the present study. 1B: Zoom-in of cluster 1 1C: Zoom-in of cluster 2.

**Figure 2:**
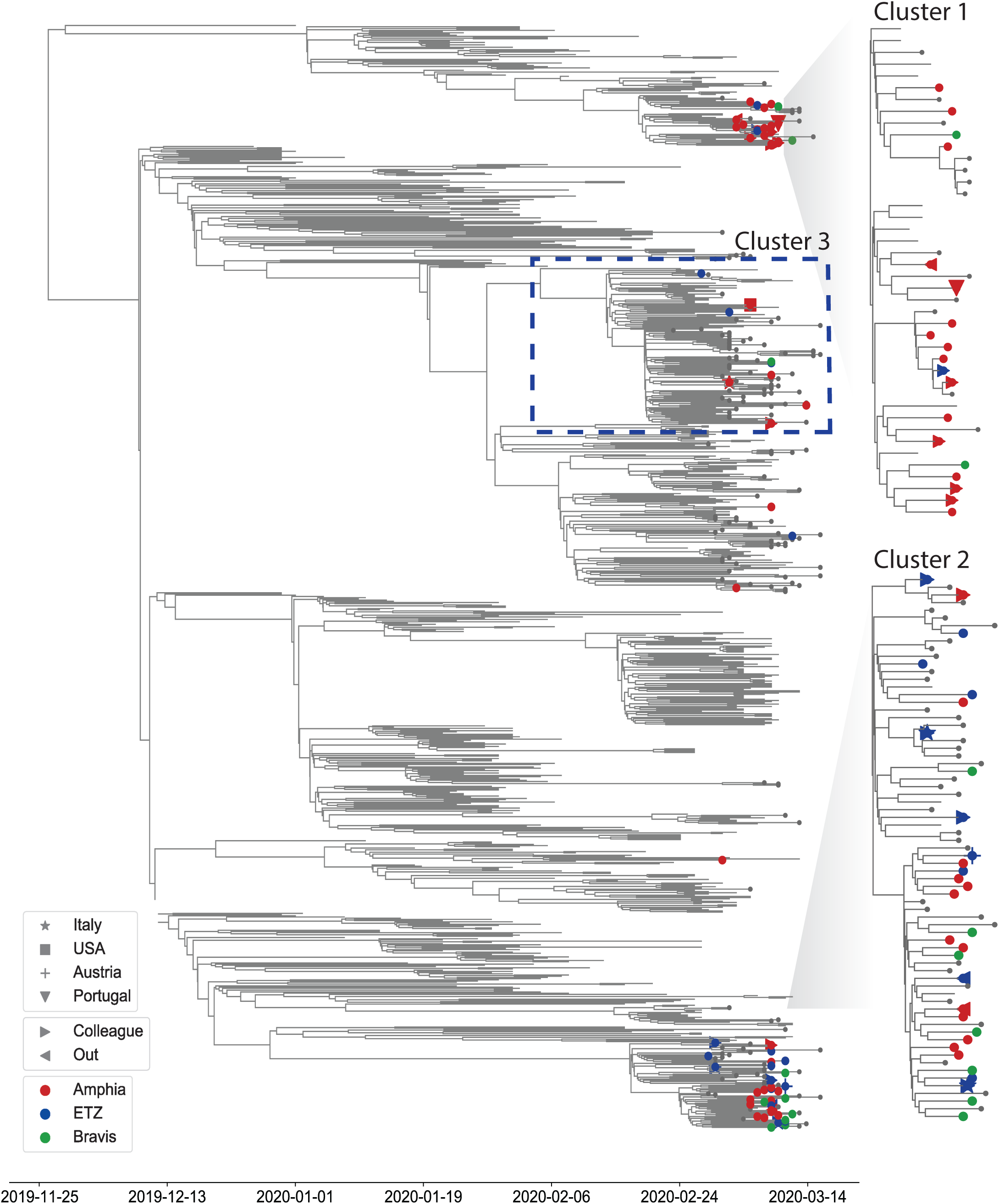
BEAST analysis of SARS-CoV-2 genomes retrieved from GISAID on March 20 2020, showing HCW contact and travel history.

Within each cluster identical or near identical sequences in HCWs in the same hospital as well as patients and HCWs were found, but there was no consistent link among HCWs on the same ward, and between HCWs and patients at the same ward. There were no clusters of more than two HCWs on the same ward with identical or near identical (1 nucleotide difference) sequences. Therefore, the observed patterns are most consistent with multiple introductions through community acquired infections, and some local amplification related to specific social events in the community. Although direct transmission in the hospitals cannot be ruled out, the data does not support widespread nosocomial transmission as source of infection in patients or healthcare workers.

## Discussion

In the present study we combined epidemiological data with whole genome sequencing to obtain a deeper understanding of SARS-CoV-2 infections in patients and staff of three hospitals in the south of the Netherlands that were the first to identify patients with COVID-19 infection in the Netherlands. Although possible hospital transmission and HCW infections with SARS-CoV-2 have been reported^3,27,28^, to our knowledge this is the first time that WGS has been used to analyze possible SARS-CoV-2 nosocomial transmission. HCW infection could have occurred through foreign travel, community contacts or nosocomial transmission. The collected epidemiological data, combined with the presence of identical viruses in all three hospitals, as well as non-hospitalized cases in other locations, indicates widespread community transmission in a very early phase of the epidemic. Mass gatherings, such as carnival, in which 63% of positive HCWs participated, possibly acted as local super spreading events.

Healthcare workers are at increased risk of being exposed to viruses within hospitals, but can also be a source of transmission by introducing a virus into their hospital. SARS-CoV-2 infections in HCWs can have a significant impact, since pathogens are introduced into settings with high numbers of individuals with co-morbidities, potentially causing high morbidity and mortality amongst patients. The current study did not find evidence of large scale nosocomial transmission, and prevailing use of PPE and other infectious disease prevention measures were considered sufficient based on these early analyses and results.

Outbreaks in healthcare settings are traditionally investigated by molecular diagnostic tools combined with epidemiological data. However, previous studies using WGS for hospital outbreak investigations have shown that the hypotheses on virus transmission routes can be incorrect based solely on these data. By adding WGS data, especially if results can be generated in a timely manner, and provided that sufficient reference sequences are available to allow a high resolution of the findings, the sequence analysis can provide essential information and inform subsequent infection control measures^29,30^.

The mutation rate of SARS-CoV-2 is estimated to be around 1.16^*^10^^^-3 substitutions/site/year, which corresponds to around one mutation per two weeks^31^. Therefore, finding identical or near identical sequences in several locations and hospitals, makes it difficult to draw definite conclusions on individual direct HCW-to-HCW or HCW-to-patient transmissions based on sequence data alone in this early stage of the SARS-CoV-2 outbreak, when the level of genetic diversity of the circulating pathogen still is limited. Moreover, we did not obtain WGS of all positive HCWs. However, the finding of diverse clusters does exclude infection from a single source. In addition, the sequence-based analysis may be biased when sampling and sequencing is not done systematically and sequence data in some areas is scarce as is the case for COVID-19 internationally. For the Netherlands, we sequenced a significant proportion of SARS-CoV-2 genomes as part of the national public health response^13^, which was used as a reference set.

We conclude that the observed genomic diversity likely reflects HCW infections contracted outside of the hospital rather than widespread within hospital transmission. Due to the near-real time sequence generation and analysis, this information was rapidly shared within the national Dutch national Outbreak Management Team. Partly based on this data, it was concluded that SARS-CoV-2 had already spread in the population in the province of Noord Brabant, which led to a change of policy, in which containment measures were complemented by targeted social distance measures, starting in the South of the country initially, and later comprising the whole country^13,32^.

## Data Availability

All data is available upon request

## Acknowledgements

We acknowledge David van der Vijver (ErasmusMC) and Harry Vennema (RIVM) for technical support.

Supplemental figure: interactive annotated Minimum Spanning Tree, displaying GISAIDname_city of residence or carnival (Prinsenbeek; Breda; Tilburg; Other)_de identified hospital wards (A to AL)_hospital

Supplemental table: GISAID acknowledgement table

